# Projecting the impact of behaviour and isolation interventions and super spreader events from mass gatherings and international travel on Malaysia’s COVID-19 epidemic trajectories using an augmented SEIR model

**DOI:** 10.1101/2020.10.29.20222224

**Authors:** Aidonna Jan Ayub, Gregory Ho Wai Son, Khayriyyah Mohd Hanafiah

## Abstract

**Background:** Various levels of lockdown implemented to contain the rapid spread of COVID-19 are not long-term solutions due to socioeconomic implications.

**Methods:** To inform safe reopening, we used an augmented SEIR model to project the impact of 1) interventions and potential new epidemic trajectories arising from super spreader (SS) events and/or international travel and 2) re-introducing strong behavioural interventions on resurgence trajectories.

**Results:** Our model suggests that 50% behaviour intervention effectiveness (BIE) (from enforced social distancing during lockdown, early in the epidemic), along with 50% isolation intervention effectiveness (IIE) (from increased testing and isolating infected individuals) was achieved during lockdown, which curbed COVID-19 transmission in Malaysia. Post-lockdown, BIE plays a minimal role if IIE reaches or exceeds 46.9% when other variables are held constant. At IIE of 30% and BIE of 21.3%, SS events of 5,000 active cases risks COVID-19 resurgence, with 4-year projected 12.9mn cumulative cases and 1.1mn deaths. Earlier action to increase BIE to 50% on day 98 compared to day 111, prevented an additional 21,401 recovered cases and 257 deaths.

**Conclusion:** Until a safe and effective vaccine is widely available, the risk of COVID-19 resurgence from large SS events warrants caution in decisions to allow for mass gatherings and regular international travel.

## 1. Introduction

The coronavirus disease 2019 (COVID-19) caused by the severe acute respiratory syndrome coronavirus 2 (SARS-CoV-2) was declared a pandemic by the World Health Organization (WHO) on 11 March 2020 [1]. The COVID-19 case fatality rate (CFR)—ranges from 0.47% to over 13% [2], and varies significantly by demographics, geographic location, and testing and healthcare capacity [3]. Compared to earlier coronavirus diseases, the Middle East respiratory syndrome (MERS) and the SARS outbreak in 2003-2004, COVID-19 is highly contagious [4,5]. Despite only emerging late in 2019, as of 22 October 2020, the COVID-19 pandemic has reached over 200 countries and territories, and over 41 million cases have been diagnosed [2].

In Malaysia, the first COVID-19 cases occurred between 24 January 2020 and 27 February 2020 amounting to 23 cases, mainly due to infected individuals arriving from overseas [6]. Community transmission of COVID-19 began on 27 February 2020, and after a number of individuals who attended an international gathering of more than 16,000 people at the Seri Petaling mosque were confirmed with COVID-19 [7], the potential scale of the epidemic in Malaysia became apparent, and the government announced a ‘movement control order’ (MCO), which for the purposes of this article is termed as a ‘lockdown’. Different iterations and phases of the lockdown have been implemented since 18 March 2020 [8], and currently the recovery phase (due to last until 31 December 2020) has significant ease in restrictions such as opening of all businesses that can adhere to social distancing and resumption of interstate travel.

Previous studies by the Malaysian Institute of Economic Research (MIER) and JP Morgan, independently estimated that Malaysia would reach a peak number of 5,070 and 6,300 active cases by mid-April, respectively [9], while another study using three different modelling methods suggested peak numbers of up to 22,000 active cases for Malaysia’s COVID-19 epidemic may be reached between mid-April and end-May [10]. In reality, Malaysia recorded a lower peak of 2,596 active cases on 5 April 2020 [2,11].Despite the unprecedented sudden disruptions in travel, economy, education and daily life for billions of people [12], the drastic social distancing measures appears to have avoided overwhelming the healthcare system and arguably, averted many preventable deaths.

Despite loosened restrictions, and reopening of domestic businesses from 13 May 2020 [8], Malaysia maintained relatively low rates of COVID-19 transmission. At the time of writing, Malaysia is experiencing a resurgence of cases, primarily within state prisons and communities in some states in the country [13]. As Malaysia and other countries continue to consider removing further restrictions, particularly regarding mass gatherings and international travel, modelling exercises can be useful to manage uncertainty and inform decisions while the pandemic continues to unfold without the availability of an effective vaccine. Thus, this study presents an augmented SEIR model to outline COVID-19 transmission trends calibrated on Malaysian data, aimed at 1) projecting transmission trends post-lockdown, to gauge potential new epidemic trajectories arising from super spreaders and/or international travel and 2) assessing the impact of re-introducing strong behavioural measures on trajectories of any subsequent epidemic waves.

## 2. Methods

### 2.1. Parameters

The model is constructed with static parameters, which are kept constant, and dynamic parameters with values that are varied throughout the period of the modelling exercise (Figure 1). Static parameters represent variables whose values do not change with time, but is based on population specific features. These include basic reproduction number (R0), incubation duration, symptomatic infection duration, and presymptomatic transmission, which were obtained based on literature review. In addition, other static parameters such as initial susceptible population, number of imported cases, isolation reaction time, fraction of COVID-19 patients needing intensive care (ICU), untreated CFR, recovery duration and duration before death; were obtained and/or estimated based on existing publicly available information. Dynamic parameters include treated CFR, testing capacity (as opposed to testing rate) (Figure S1), critical care capacity (Figure S2), and super spreaders, which were based on reports from government agencies and secondary sources, and behaviour intervention effectiveness (BIE), and isolation intervention effectiveness (IIE), which were determined by authors for scenario projection purposes (Table S1).

**Figure 1.**
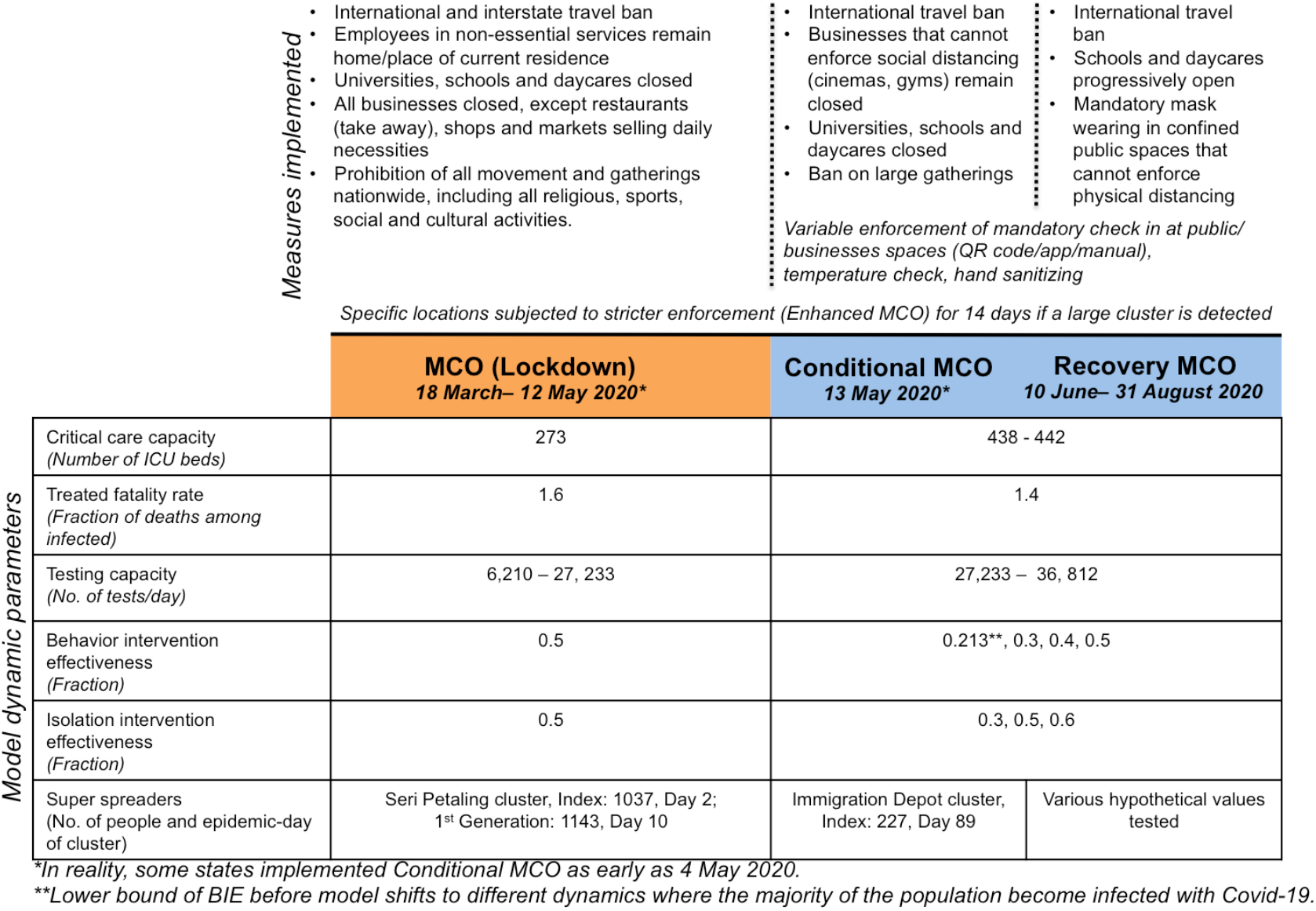
Model dynamic parameters over different phases of lockdown, constructed with reference to [8]

**Figure 2.**
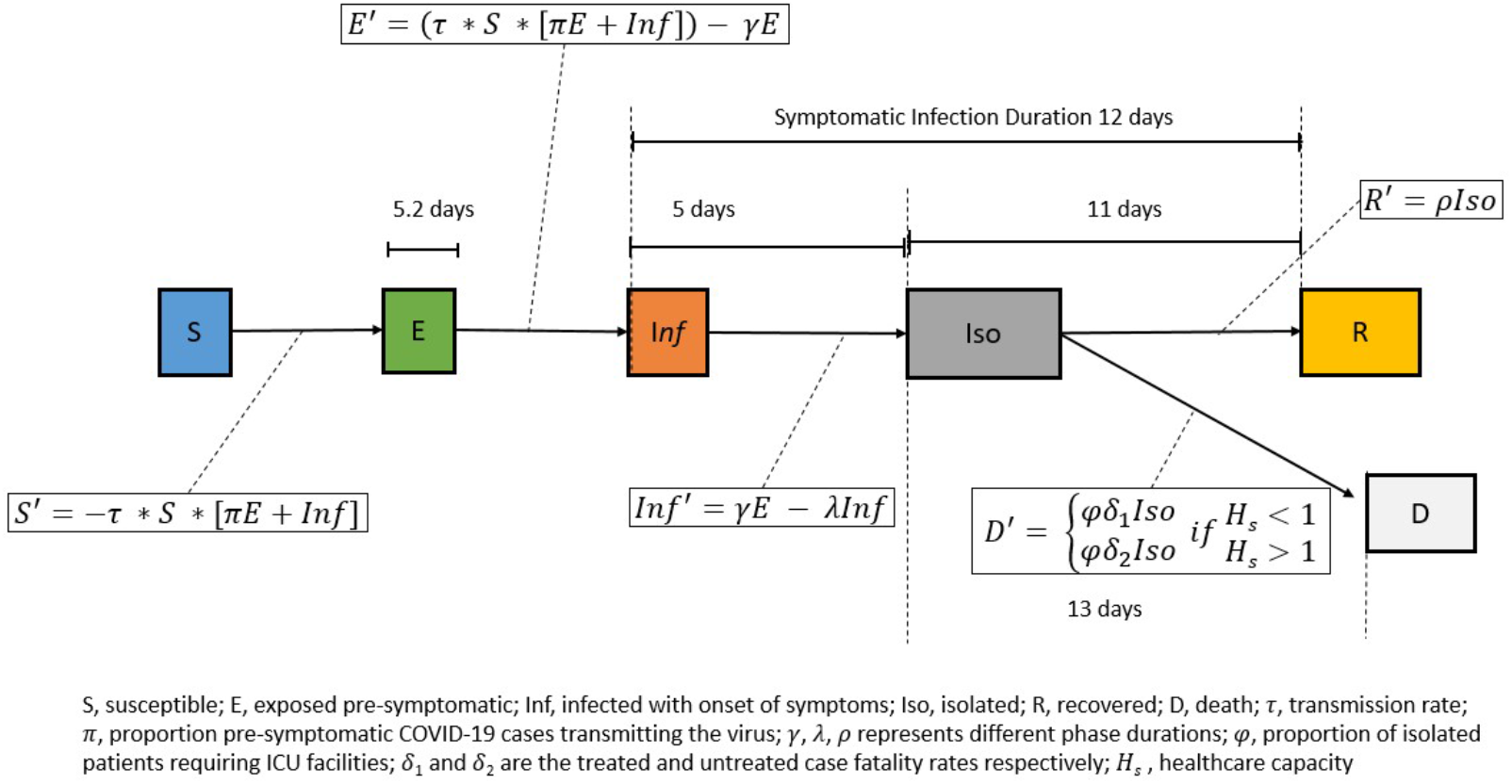
Compartments of augmented SEIR model.

Specifically, we attempt to model the scale of change arising from interventions and SS events on population transmission dynamics. For the purpose of this analysis, BIE reflects the effect of practices such as social/physical distancing (with the strictest version of distancing observed under lockdown), wearing masks, and frequent hand washing; IIE reflects testing, contact tracing and isolation of infected persons. Both parameters of effectiveness are constructed using a proportional effectiveness scale of 0% (least effective) to 100% (fully effective). Additionally, although various definitions exist for the term super spreader(s) cluster/event/episode [14], here we define ‘super spreaders (SS)’ as a typically large cluster of active cases within one to two infection generations that unpredictably arise due to various biological, host and environmental factors such as large congregations or from imported cases i.e. international arrivals who test positive for COVID-19 within quarantine period [15]. We also do not model the transmission dynamics within SS clusters or events themselves.

### 2.2 Model features and key assumptions

This study uses an SEIR model (Figure 1), built using System Dynamics on Vensim by Ventana Systems, adopted from a basic model built by Tom Fiddaman of Ventana Systems [16]. The model was augmented to include presymptomatic transmission, SS events, testing capacity, a scaling up of critical care beds, BIE and IIE set at different levels pre- and post-lockdown, and BIE set at different levels to allow for simulating earlier and later action (Figure S3). In addition, the density of social networks was removed due to data limitations while the impact of seasons was removed, as Malaysia is a tropical country.

Key assumptions of the model include: 1) homogeneous mixing, whereby all individuals have equal characteristics, and equal susceptibility to infection, morbidity and mortality, despite the fact that age-structure, comorbidity prevalence, and differences due to social structures or geographies in the population are known to influence disease spread and outcomes [3]; 2) all active cases are isolated through hospitalization, and there are no COVID-19 re-infections or waning immunity, despite some reports suggesting reinfections are possible [17]; 3) asymptomatic (as opposed to presymptomatic) cases do not influence transmission, as it remains unclear to what extent they contribute to the spread of the disease [18–20]; 4) R0 is 3.9, based on average Malaysian household size of 3.9 [21,22]; 5) hospital strain is defined as the fraction of severe cases requiring care at the intensive care unit (ICU), estimated by the number of ICU beds in COVID-19 admitting public hospitals and used to determine CFR, whereby when ICU capacity is exceeded, the CFR would converge at a higher level where ICU treatment becomes unavailable as was seen in countries like Italy and Spain [2] and 6) effectiveness of interventions, specifically BIE from implementation of and adherence to the lockdown was constant over specific periods of time (i.e. during and post-strict lockdown), although in reality there were different levels of movement restriction and compliance at different times.

### 2.3 Model testing and projections of super spreader events

Sensitivity analysis was conducted on the final model to assess robustness and identify parameters and parameter values that have the highest impact on model trends. The baseline model was then benchmarked against publicly available data from the Ministry of Health (MOH) Malaysia [2,11].

To project scenarios of potential resurgence, the model was seeded with various sizes of SS events on day 77 since community transmission began (13 May 2020, post-strict lockdown), while alternating IIE and BIE at different levels, to observe the impact on COVID-19 trajectories over a 4-year period.

Then, to observe the impact of strong interventions such as lockdowns on a subsequent COVID-19 wave, the model was seeded with various sizes of SS events on day 89 since community transmission began (25 May 2020). Day 89 was chosen for this analysis because this was when Malaysia experienced an uptick in cases due to immigration detention depot clusters, thus providing a counterfactual to our model results. A higher BIE (lasting for 14 days) was then introduced at an earlier timepoint (day 98) and a later timepoint (day 111) post-lockdown.

## 3 Results and Discussion

### 3.1 Model calibration to Malaysian COVID-19 epidemic curve suggests effectiveness of behaviour intervention

After calibration, the trend of active cases and deaths projected by the augmented SEIR model was comparable to Malaysia’s epidemic curve, with minor exceptions. The model’s projected peak was 2,630 isolated cases on 29 March 2020 and cumulative number of deaths at 166 cases, while Malaysia recorded a peak of 2,596 active cases on 5 April 2020 and cumulative number of deaths at 125 cases as of 17 August 2020 (Figure 3a). The data suggests that the implementation of behaviour interventions, specifically the lockdown, had a significant impact in reducing the peak and increasing the rate of declining cases. This appears in line with the effect of lockdowns to suppress the initial wave of COVID-19 community transmissions taken by countries such as China [23,24], and even on subsequent waves such as in Victoria, Australia [25], specifically international travel restrictions and workplace closure, which appears to reduce the R0 [26], rapidly bringing down the number of new and active cases of COVID-19, and reducing mortality [27].

**Figure 3.**
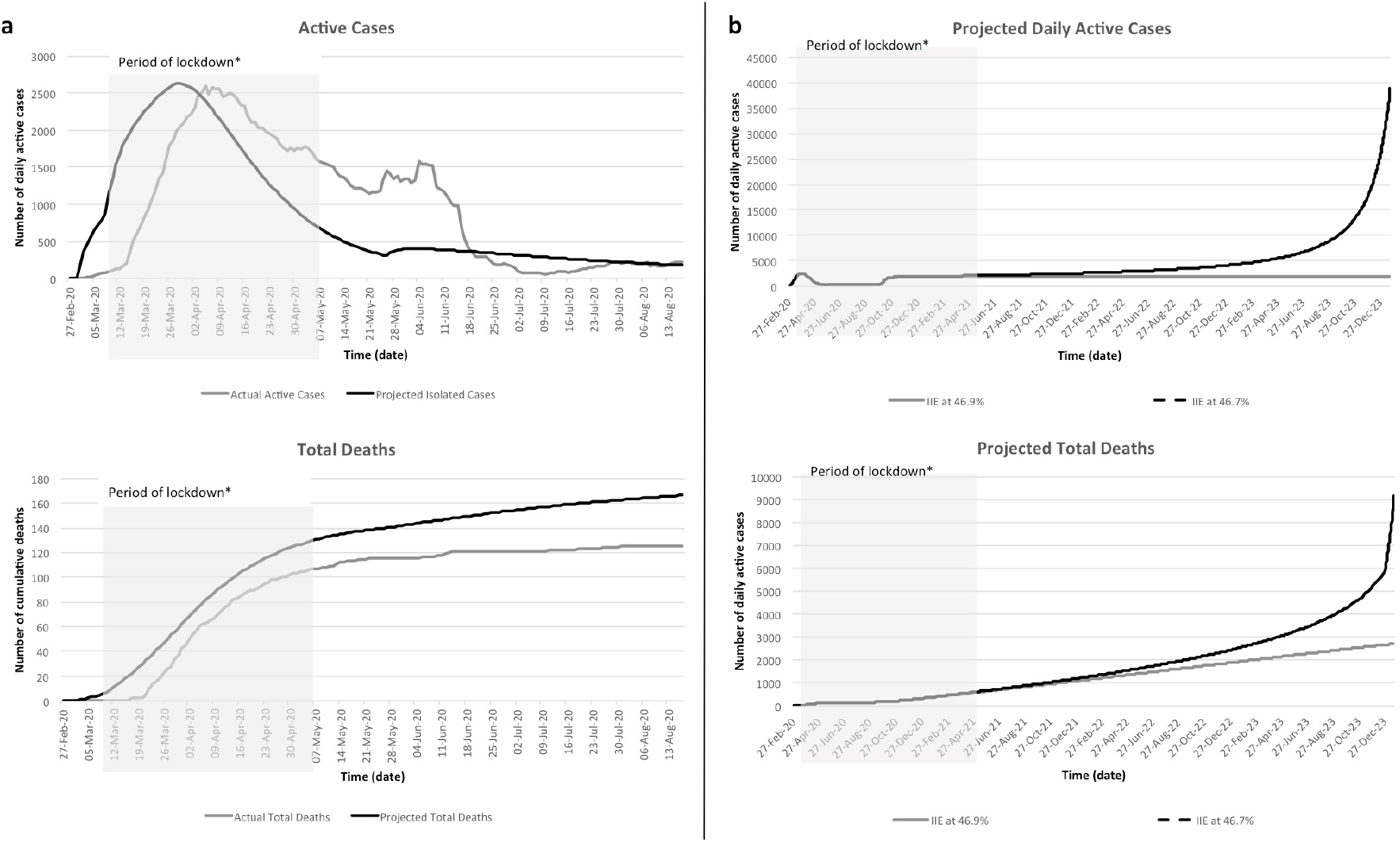
a) Calibration of model at baseline compared to Malaysian MOH data on active cases and total deaths; b) Projected daily active cases and deaths over 4-years at threshold, and below threshold IIE when BIE is at 0%. Note: *Some states had started to ease restrictions as early as 1 May 2020.

**Figure 4.**
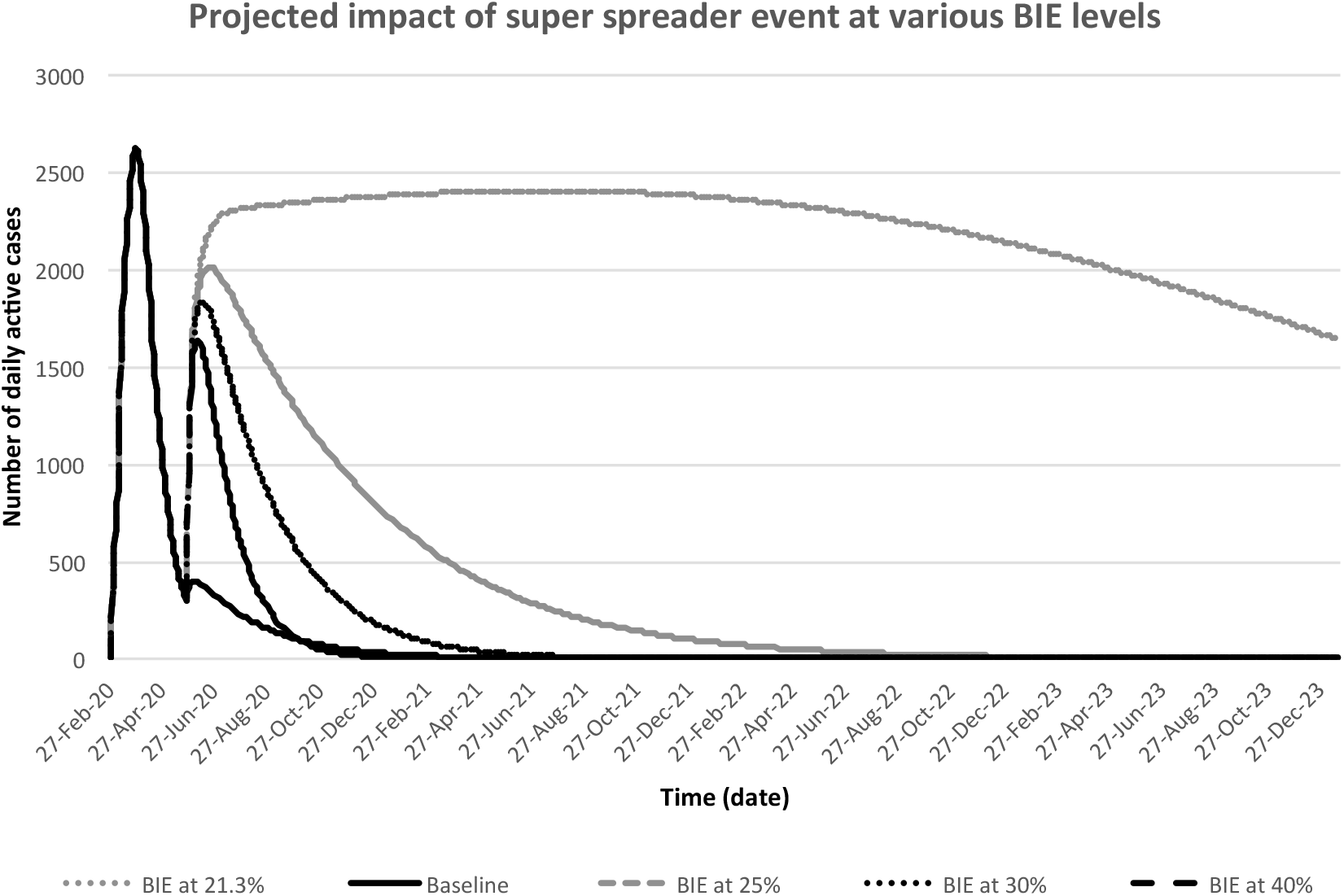
Projected impact of super spreader event size of 2,000 cases (at single seeding event) at IIE of 30% and different levels of BIE on COVID-19 trajectories post-lockdown.

The calibration of the baseline model to the data further suggest that isolation and behaviour interventions likely achieved up to 50% effectiveness during the lockdown. However, these numbers do not reflect specific interventions, and they are not comparable numeric measurements of effectiveness. Instead IIE and BIE represent the summative effect of various actions that reduce the risk of infected persons from spreading the disease, and uninfected persons from being exposed to the disease, respectively.

Furthermore, the model suggests that when IIE exceeds 46.9%, the number of deaths converge sub-linearly at the treated CFR of 1.7% (Figure 3b), and at higher levels of IIE, changes to the BIE has minimal influence on projections of active cases or deaths. Conversely, reducing IIE below 46.9% while holding the BIE at 0% appears to push the model into different dynamics where a majority of the population become infected with COVID-19, and the trend of active, recovered and deaths scale super-linearly at the untreated CFR. This underscores the idea that testing and isolation should remain the key strategies in controlling COVID-19.

Additionally, maintaining sufficiently high levels of IIE reduces risk of resurgence post-lockdown even at minimum BIE. This perhaps reflects the ability of countries such as South Korea and Taiwan which have been able to control COVID-19 primarily through ramping up early testing, contact tracing and isolation relative to the size of its outbreak, without undergoing a full lockdown [28,29]. However, the importance of strong behavioural interventions early in the epidemic to flatten the curve cannot be discounted as the enforcement of strict border controls and lockdown early are known to have played a crucial role in controlling transmissions as seen in China and a number of countries across sub-Saharan Africa [30–32].

One caveat, however, is the fact that IIE and BIE are treated as separate parameters in the model. This neglects the reality that restrictions in civilian movement likely influence the effectiveness of contact tracing and isolation. Additionally, although BIE was likely not constant, we deduced it reached 50% once compliance to lockdown was reportedly 95%, based on model calibration exercises [33], and was maintained at 50% until the end of strict lockdown. And although IIE was likely not constant and much lower than 50% at the beginning of lockdown compared to its end, we tried to account for this by having testing capacity scaled over time to reach max capacity from 6,210 tests – 36,812 tests.

Highlighting a clear limitation of the homogeneity assumption in SEIR modelling, the model was unable to sufficiently project the effects of a second lower peak on 6 June 2020, which arose from outbreaks among mainly undocumented foreign workers at a number of immigration detention depots in the country. As the outbreaks occurred in populations that were significantly isolated from the larger community, it is likely that the rapid rise in new transmissions was able to be controlled much faster using intensified targeted approaches, resulting in the rapid drop to less than 1000 active cases by 14 June 2020 [34]. Additionally, the model was unable to account for a change in patient discharge criteria, based on an updated WHO recommendation to release patients from isolation without requiring re-testing published on 27 May 2020 [35], which resulted in a rapid decline in active cases after 12 June 2020 [36].

### 3.2 At minimum isolation and behaviour intervention effectiveness, a super spreader cluster above 2,000 cases risks COVID-19 resurgence and overwhelming the healthcare system

The second peak due to outbreaks across immigration detention depots highlighted the potential for new clusters or super spreaders that may occur to drive new waves of COVID-19 in the country, particularly with further ease in restrictions and changes to policies on mass gatherings, and policies on international travel and/or on quarantine of overseas travellers. Additionally, several studies increasingly highlight the role of SS events, particularly those arising from social settings, in driving the bulk of transmissions [37,38]. Hence, we attempted to model these potential super spreaders of varying sizes, under scenarios of varying levels of BIE (with IIE held constant at 30%), and their potential to cause a subsequent wave that may warrant strong measures to avoid overwhelming the health system.

Based on this analysis, SS events of up to 2,000 new active cases may still be manageable by the current health system, with the 4-year projected number of recovered (cumulative number of infected patients who survived) varying from 19,425 to 273,953 people and deaths varying from 243 to 3,301 people, assuming that BIE is between 40% and 21.3%. Beyond these numbers, a SS event size of 5,000 or 26,000, assuming that BIE at the time ranged from 21.3% to 40%, results in 136,342 recovered and 1,648 death cases or 12,924,200 recovered and 1,097,860 death cases, respectively (Table 1). Above the threshold SS event size or above 2,000 cases under Malaysia’s current scenario, the BIE has significant influence on the long-term COVID-19 trajectory. Specifically, at BIE of 40%, a super spreader cluster size of 26,000 cases results in a CFR of 1.2%, while at BIE of 21.3%, a SS event size of 5,000 cases results in a higher CFR of 7.8%.

**Table 1.** The impact of super spreader (SS) event size on Covid-19 projected recovered cases and deaths based on different levels of behavior intervention effectiveness (BIE)

| Projection | BIE (%) | SS event size | 4 year-cumulative |  |  |
| --- | --- | --- | --- | --- | --- |
|  |  |  | Recovered | Dead | CFR** (%) |
| **Baseline** | **30.0** | **227** | **14,321** | **182** | **1.3%** |
| Risky Zone | 40.0 | 26,000 | 136,342 | 1,648 | 1.2% |
| Safer Zone | 40.0 | 2,000 | 19,425 | 243 | 1.2% |
| Risky Zone | 30.0 | 26,000 | 400,923 | 4,826 | 1.2% |
| Safer Zone | 30.0 | 2,000 | 28,714 | 354 | 1.2% |
| Risky Zone | 25.0 | 17,000 | 9,410,680 | 740,330 | 7.3% |
| Safer Zone | 25.0 | 2,000 | 50,801 | 620 | 1.2% |
| Risky Zone | 21.3* | 5,000 | 12,924,200 | 1,097,860 | 7.8% |
| Safer Zone | 21.3* | 2,000 | 273,953 | 3,301 | 1.2% |

Notes: \*21.3% is determined to be lower bound of BIE before model shifts to dynamics where a majority of the population becomes infected with COVID-19. Range of treated.\*\* Range of treated CFR for Malaysia is 0-1.7% while untreated CFR is assumed to be 10%.

Given the socioeconomic costs of lockdowns and difficulty to maintain compliance with social distancing and required self-isolation over longer periods of time [39], countries may prevent resurgence by avoiding situations that enable surpassing the threshold super spreader numbers, which may arise from large gatherings and/or imported cases. To date, policies in managing the risk of imported cases, which include mandatory quarantine at government approved lodging and screening tests appear to be effective in managing the spread of the disease from import cases. Thus far other countries that have maintained strict border control and/or strict quarantine such as Singapore, New Zealand, South Korea and Hong Kong have avoided significant resurgence in cases, while other countries such as the UK, Spain and Germany with less rigorous testing and control of influx of international travellers appear to be having rapid increase in cases post-lockdown [40]. That said, such measures are likely insufficient to address border entries through undocumented channels, which is compounded with difficulty to detect infected individuals until an outbreak becomes more apparent as is the case with Malaysia.

Similarly, large conferences, and other mass gatherings including regular religious and community events such as congregational prayers and weddings that may garner up to 1,000 to tens of thousands of attendees, pose significant risk for resurgence. In particular, large events that host international and/or interstate participants increases mixing of previously separated people carrying different strains of the virus [41], raising the risk of sparking SS events and dispersing new clusters, which may spark another large COVID-19 wave that may overwhelm the health system. Such risks are heightened by the possibility of presymptomatic transmissions from COVID-19 infected persons as early as 3 days before the appearance of symptoms [20,42,43], and the increasing burden and decreasing effectiveness in tracing very large groups of contacts [29,44].

Conversely, the model also suggests that the threshold super spreader cluster size that remains manageable by the health system can be as high as 26,000 cases, depending on what is viewed as an acceptable range of recovered and death cases, what is the presumed BIE level (perhaps due to strict observance of physical distancing and mask wearing) at a particular period, and whether the number of critical care beds could be increased. Regardless, until an effective vaccine becomes available, mass gatherings, international travel and re-opening of borders require careful deliberation and targeted policies.

### 3.3. Impact of strong and earlier action on trajectories in the event of COVID-19 resurgence

Many countries imposed lockdowns to ‘flatten the curve’, i.e. to prevent a large number of new infections and spread them over a longer period of time to allow healthcare systems to cope with influx of COVID-19 patients, and prevent fatalities that would otherwise occur due to an overwhelmed healthcare system. However, while the baseline model calibrated to Malaysian COVID-19 data shows the positive impact of the lockdown on reducing COVID-19 transmission, the restrictions imposed have significant socioeconomic costs [45,46] and ramifications on mental health and wellbeing for societies at large [47]. Thus, although governments may be cautious to re-introduce a lockdown after reopening and achieving low rates of COVID-19 transmission, this modelling exercise reaffirms the need for implementing strong actions such as lockdowns and doing so earlier rather than later should a subsequent COVID-19 wave arise from large super spreaders.

In the event of a COVID-19 resurgence, whereby BIE is assumed to be 21.3%, implementing the lockdown reduces cumulative cases by 13.5 million people, and deaths by 1.1 million people across a 4-year period (Table 2), with projections varying according to BIE.

**Table 2.** The impact of strong action (lockdown) to increase behavior intervention effectiveness (BIE) on COVID-19 trajectories post-super spreader (SS) events and difference between earlier and later intervention

| BIE (%) | SS event size | *No Lockdown* |  |  | *Earlier Lockdown*<br>*(9.2 days)* |  | *Later Lockdown*<br>*(22.2 days)* |  | *Difference*<br>*(Earlier – Later)* |  |  |
| --- | --- | --- | --- | --- | --- | --- | --- | --- | --- | --- | --- |
|  |  | Additional Positive | Additional Recovered | Additional Dead | Recovered | Dead | Recovered | Dead | Additional Cases | Additional Recovered | Additional Dead |
| 21.3* | 5,000 | 13,485,092 | 12,393,616 | 1,091,476 | 530,584 | 6,384 | 551,985 | 6,641 | 21,658 | 21,401 | 257 |
| 25 | 17,000 | 9,785,442 | 9,049,461 | 735,981 | 361,219 | 4,349 | 388,520 | 4,677 | 27,629 | 27,301 | 328 |
| 30 | 26,000 | 163,714 | 161,770 | 1,944 | 239,153 | 2,883 | 257,295 | 3,101 | 18,360 | 18,142 | 218 |
| 40 | 26,000 | 17,704 | 17,494 | 210 | 118,848 | 1,437 | 123,872 | 1,498 | 5,084 | 5,024 | 60 |

\*Notes: 21.3% is determined to be lower bound of BIE before model shifts to dynamics where a majority of the population becomes infected with COVID-19.

However, a key issue in implementing a lockdown is the level of enforcement and timing, and early strong action has been argued to have more significant impact compared to delayed or ineffective action [48]. In this model, we assume that early strong action equals to imposing a strict lockdown 9.2 days after index case in a super spreader event is determined, while delayed action equals to 22.2 days, or 13 days later. Delay in taking strong actions may cost between 5,084 and 27,629 additional COVID-19 cases and an additional 60 to 328 deaths, depending on the size of the super spreader and assumed levels of BIE.

While there appears to be discrepancy of numbers of active/isolated cases and deaths projected for earlier versus later lockdown implementation compared to the numbers of active/isolated cases and deaths projected for lockdown versus no lockdown, the former may be a more reliable projection. Modelling scenarios of a lockdown versus no lockdown does not consider timepoint of action, and only looks at the cumulative impact in four years, which is arguably more susceptible to various unpredictable factors over time, and unlikely to be observed given that governments are likely to take mitigatory action. Comparing earlier versus later implementation of a lockdown relative to timepoint along the epidemic curve, may be more useful as it illustrates when the decision to intervene may change trajectories, and to what extent.

Nevertheless, the purpose of this exercise is not to predict or estimate numbers of cases, particularly on the difference observed when projections made at behaviour intervention effectiveness of 21.3% (∼13mn) and or 40% (∼17k). Instead, we aim to show the scale of change on the trajectories depending on the time of the decision point to enforce strong actions such as lockdowns. While we explored the impact of timing of strong actions to mitigate resurgence post-lockdown, our findings also reiterate the point that earlier action may have played an even bigger role earlier in the epidemic. This is because enforcing lockdown early enough in the epidemic to increase BIE, provides the opportunity for IIE to increase (which requires time to accumulate sufficient knowledge, resources and capacity). Because COVID-19 is now known to be highly overdispersed, in which a small fraction of infected people could be responsible for approximately 80% of transmissions [38], high IIE post-lockdown halts infected persons with high potential for sparking super spreader clusters and controls the epidemic more effectively compared to BIE. This also enables countries to reduce restrictions on general population behaviour, allowing the resumption of domestic mobility and socioeconomic activities without necessarily causing a subsequent COVID-19 wave, as long as effective testing and isolation strategies are maintained, and situations that exceed the threshold cluster sizes are avoided.

That said the model is limited in its ability to capture the potential influence of changing trajectories of COVID-19 on both BIE and IIE. For example, while there may be a scenario where transmissions post 26,000-case super spreaders remains within SEIR dynamics at IIE of 30% and BIE of 40%, arguably at such high levels of community transmission, the effectiveness of these interventions will likely be reduced. Active case numbers post-lockdown in the model is much lower than what is recorded by the Malaysian MOH, which would affect the impact of the IIE and BIE values on scenario projections. These caveats necessitate more conservative interpretations and framing discussions on trends of COVID-19 arising from super spreaders and consequent actions taken to mitigate these based on the projections of additional positive and death cases at the lower bound of 21.3% BIE.

Other key limitations in this modelling exercise include 1) heterogeneity factors such as age, comorbidities, and social structure and geographies, and economic implications of interventions, were not included in the model; 2) whether the index case in an SS event is from local transmission or an imported case (potentially carrying different strains of the virus with different virulence and transmission capability [41] is not directly considered. In particular, the variation in propensity for different individuals to spread and disperse disease or spark new clusters, and poor characterisation of the role of typically asymptomatic children in transmitting disease [3], are significant limitations in projecting longer-term disease trends. Ultimately, the model could be improved with more granular information on the local dynamics of the disease, transmission, and healthcare system.

## 4. Conclusions

This study attempted to model the longer-term impact of COVID-19 super spreaders that may potentially arise from mass congregations and international travel, and the value of strong early action, such as imposing lockdown, on changing epidemic trajectories. While the model presented here has been contextualized to the Malaysian COVID-19 epidemic, our analysis is well-aligned with others that have previously reported the pivotal role of testing and isolation strategies and the importance of maintaining high levels of their effectiveness. The recent resurgence of COVID-19 in several cities that had previously contained the spread of the virus reiterates the importance of managing potential sizes of super spreaders that may result in subsequent epidemic waves, and the value of early action. As countries continue to consider safe strategies of re-opening and easing restrictions in the absence of accessible, safe and effective vaccines, the findings of this study argue for maintaining caution in decisions pertaining to mass gatherings and/or regular international travel to avoid enabling large super spreader clusters of COVID-19 cases to arise, which may necessitate re-imposition of stronger and socioeconomically costly action such as lockdowns.

## Data Availability

Data is available upon request.

## Conflicts of Interest

The authors declare no conflicts of interest.

## Acknowledgements

The authors would like to thank Adamzairi Bahaud Din, Muhammad Hafiz Wan Rosli, and Lai Kah Chun for their input to the study, and Kathryn Jacobsen and Illisriyani Ismail for reviewing earlier versions of the manuscript and model, and Khazanah Research Institute for financial and operational support. The views and opinions expressed are those of the author and may not necessarily represent the official views of Khazanah Research Institute. All errors remain authors’ own.

## Funding

The authors did not receive specific funding for this work. The authors would like to thank Khazanah Research Institute for allowing the authors to apportion time for this work.

## Notes

### Competing Interest Statement

The authors have declared no competing interest.

### Author Declarations

This study does not involve direct human subjects and relies on secondary publicly available data.

