## Supplemental Material for "Projecting the impact of behaviour and isolation interventions and super spreader events from mass gatherings and international travel on Malaysia’s COVID-19 epidemic trajectories using an augmented SEIR model"

**Supplementary Materials**

Table S1. Parameter and parameter values input to the model

| **Input Parameters** | **Definition** | **Input Values** | **Range of Value(s)** | **Reference(s)** |
|---|---|---|---|---|
| Initial Susceptible Population | Malaysian population as of 2019 | 32.68 | 32.68 million people | DOSM (2019)[1] |
| Number of imported cases | Total number of positive cases at the start of community transmission | 23* | 23 people | MOH (2020)[2] |
| Basic reproduction number (R0) | Epidemiologic metric to indicate contagiousness | 3.9 | 1.5 – 4.5 | Delamater et al. (2019)[3]; F. He et al., (2020)[4]; DOSM (2019)[1] |
| Incubation Duration | Time between exposure to virus (becoming infected) and onset of symptoms | 5.2 | 5 –14 days | WHO (2020)[5]; ASM (2020)[6] |
| Symptomatic Infection Duration | Duration from time of symptoms appearance/ diagnosis to recovery or death | 12 | 4 - 17 days | Azrai et al., (2020)[7] |
| Isolation Reaction Time | The time it takes to diagnose and isolate/ quarantine/hospitalise an infected person | 5 | 2 - 11 days | ASM (2020)[6]; Azrai et al., (2020)[7] |
| Testing Capacity | Average number of tests completed per day | Increasing scale from 6,210 to 27,233 to 36,812 | 6,210 - 36,812 tests per day | MOH (2020) [8] |
| Behaviour Reaction Time | The time point when lockdown was implemented with high compliance (relative to the start of community transmission) | Day 25 | Day 25 (22 March 2020) | PMO (2020) [9] |
| Presymptomatic Transmission | Proportion of transmissions from infected people who have not developed symptoms | 12.6%** | 6.4%, 12.6%, 44% | Wei et al. (2020); X. He et al. (2020); Kimball et al. (2020)[10–12] |
| Presymptomatic Duration | Days patient is shedding/ spreading the virus before showing symptoms | 3 | 1-3 days | Wei et al. (2020); X. He et al. (2020); WHO (2020)[10,11] |
| Critical Care Capacity | Number of ICU beds in public hospitals admitting COVID-19 patients | Fixed increase from 273 to 438 to 442 | 273 - 442 beds | MOH, 2020 [8] |
| Fraction needing ICU | Proportion of hospitalised infected people who require ICU. Estimated based on data up to 17 August 2020 | 1.70% | 0.0 – 7.2% | Estimated from Azrai, Yue Lok, and Suait (2020) and Outbreak.my [7,13] |

| Input Parameters | Definition | Input Values | Range of Value(s) | Reference(s) |
|---|---|---|---|---|
| Treated Case Fatality Rate | The proportion of infected people who receive medical care/hospitalisation (currently, all cases) and die from COVID-19. Estimated based on data up to 17 August 2020 | 1.6% (pre/during lockdown), 1.4% (post lockdown) | 0 - 1.7% | Outbreak.my [13] |
| Untreated Case Fatality Rate | The proportion of infected people who do not receive medical care/hospitalisation and die from COVID-19 | 10%*** | 10 – 13% | Case fatality rate of countries with surged hospital capacity (Spain and Italy) Outbreak.my [13] |
| Recovery Duration | Number of days between diagnosis and recovery | 11 | 4 - 21 days | Estimated from Outbreak.my [13] |
| Duration Before Death | Number of days between symptoms appearance and death | 13 | 0 – 87 days | Estimated from Azrai et al., (2020)[7] |
| Super spreader (Seri Petaling) cluster | Large cluster of infected people. The first large cluster in Malaysia is known as the Seri Petaling cluster. Number of cases include index cases and first generation cases. | 1,037 (index cases) on model's Day 2 and 1,143 (first generation cases) on model's Day 10 | 2,180 people | MOH (2020)[14] |
| Isolation Intervention Effectiveness | Variable reflecting the effectiveness of isolation, testing and contact tracing by the public health and healthcare systems | Differs according to scenario | 0.0 - 0.5 | Author’s estimation |
| Behaviour Intervention Effectiveness | Variable representing the effectiveness of behavioural interventionsions such as social distancing, washing hands and wearing masks | Differs according to scenario | 0.0 – 0.5 | Author’s estimation |
| Post-lockdown day | The time since start of community transmission to when strict lockdown ends | Day 77 | Day 77 (13 May 2020) | Author’s calculation |

Note: Estimates are based on publicly available information.
* Although not all 23 cases were imported cases, for the purposes of simplifying our model, we assume that they are all imported cases.
** Presymptomatic transmissions have been documented. However, the proportion of infected persons who are presymptomatic and spreading the disease is less clear. For this model, 12.6% is the proportion of presymptomatics reported by (Kimball et al. 2020), although the study was not able to confirm whether all 12.6% were also spreading the disease.

*** In the model, the value differs according to whether there is hospital strain. This number is estimated based on the fatality rate of countries that had apparently exceeded health system capacity due to COVID-19.

[1] Department of Statistics Malaysia Official Portal. Demographic Statistics Fourth Quarter 2019, Malaysia.

https://www.dosm.gov.my/v1/index.php?r=column/cthemeByCat&cat=430&bul_id=UDc0eVJ4WEJiYmw0Rmt5cjYvWHFkdz09&menu_id=L0pheU43NWJwRWVSZklWdzQ4TlhUUT09 [accessed 25 March 2020].
[2] Kenyataan Akhbar KPK 29 Februari 2020: Situasi Semasa Jangkitan Penyakit Coronavirus 2019 (COVID-19) di Malaysia 2020. https://kpkesihatan.com/2020/02/29/kenyataan-akhbar-kpk-29-februari-situasi-semasa-jangkitan-penyakit-coronavirus-2019-covid-19-di-malaysia/ [accessed 22 April 2020].
[3] Delamater PL, Street EJ, Leslie TF, Yang YT, Jacobsen KH. Complexity of the Basic Reproduction Number (R0). Emerg Infect Dis 2019;25:1–4. https://doi.org/10.3201/eid2501.171901.
[4] He F, Deng Y, Li W. Coronavirus Disease 2019: What We Know? J Med Virol 2020;n/a. https://doi.org/10.1002/jmv.25766.
[5] Coronavirus Disease 2019 (COVID-19) Situation Report – 73. 2020.
[6] Academy of Sciences Malaysia. COVID-19 RESEARCH STUDIES FACT SHEET 1 2020. https://www.akademisains.gov.my/covid-19/
[7] Azrai E, Yue Lok C, Suait S. Covid19-KKM. Google Sheet: 2020. https://docs.google.com/spreadsheets/d/15A43eb68LT7gg_k9VavYwS1R2KkCTpvigYMn5KT9RTM/edit#gid=41230890
[8] Kenyataan Akhbar KPK 25 Jun 2020 – Situasi Semasa Jangkitan Penyakit Coronavirus 2019 (COVID-19) di Malaysia 2020. https://kpkesihatan.com/2020/06/25/kenyataan-akhbar-kpk-25-jun-2020-situasi-semasa-jangkitan-penyakit-coronavirus-2019-covid-19-di-malaysia/ [accessed 25 June 2020]
[9] Perutusan Khas YAB Perdana Menteri (25 Mac 2020) 2020. https://www.pmo.gov.my/2020/03/perutusan-khas-yab-perdana-menteri-25-mac-2020/ [accessed 24 April 2020].
[10] Wei WE, Li Z, Chiew CJ, Yong SE, Toh MP, Lee VJ. Presymptomatic Transmission of SARS-CoV-2 — Singapore, January 23–March 16, 2020. Morb Mortal Wkly Rep 2020;69:411–5. https://doi.org/10.15585/mmwr.mm6914e1.
[11] He X, Lau EHY, Wu P, Deng X, Wang J, Hao X, et al. Temporal Dynamics in Viral Shedding and Transmissibility of COVID-19. Nat Med 2020. https://doi.org/10.1038/s41591-020-0869-5.
[12] Kimball A, Hatfield KM, Arons M, James A, Taylor J, Spicer K, et al. Asymptomatic and Presymptomatic SARS-CoV-2 Infections in Eesidents of a Long-Term Care Skilled Nursing Facility — King County, Washington, March 2020. MMWR Morb Mortal Wkly Rep 2020;69:377–81. https://doi.org/10.15585/mmwr.mm6913e1.
[13] COVID-19 | Malaysia Outbreak Monitor | Live Updates n.d. https://www.outbreak.my.
[14] Kenyataan Akhbar KPK 8 Julai 2020 – Situasi Semasa Jangkitan Penyakit Coronavirus 2019 (COVID-19) di Malaysia 2020. https://kpkesihatan.com/2020/07/08/kenyataan-akhbar-kpk-8-julai-2020-situasi-semasa-jangkitan-penyakit-coronavirus-2019-covid-19-di-malaysia/ [accessed 8 July 2020]

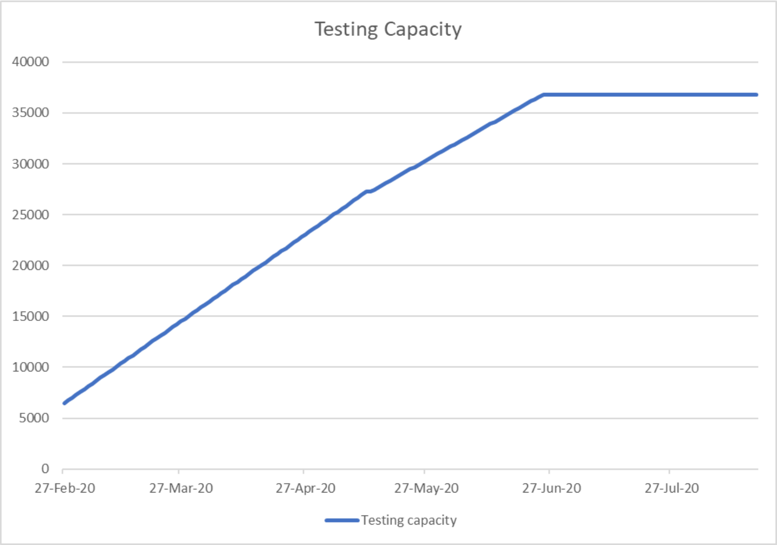


Figure S1. Testing capacity. The line graph shows scaling up of testing capacity over time taken into account in the model.

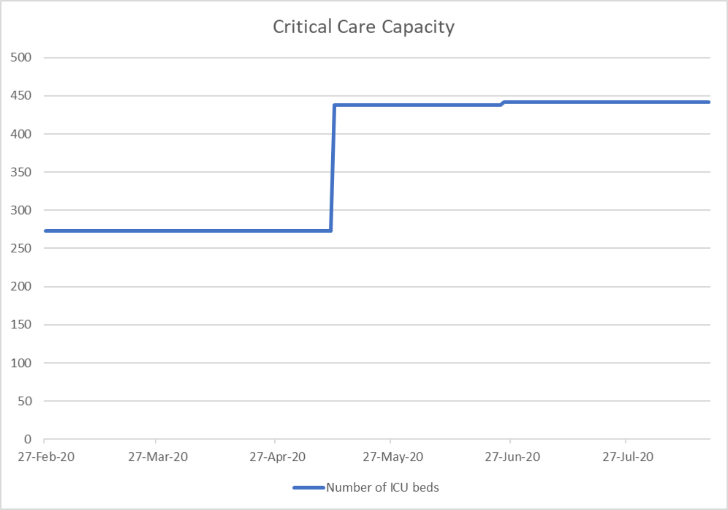


Figure S2. Critical care capacity. The graph shows change in critical care capacity over time taken into account in the model.

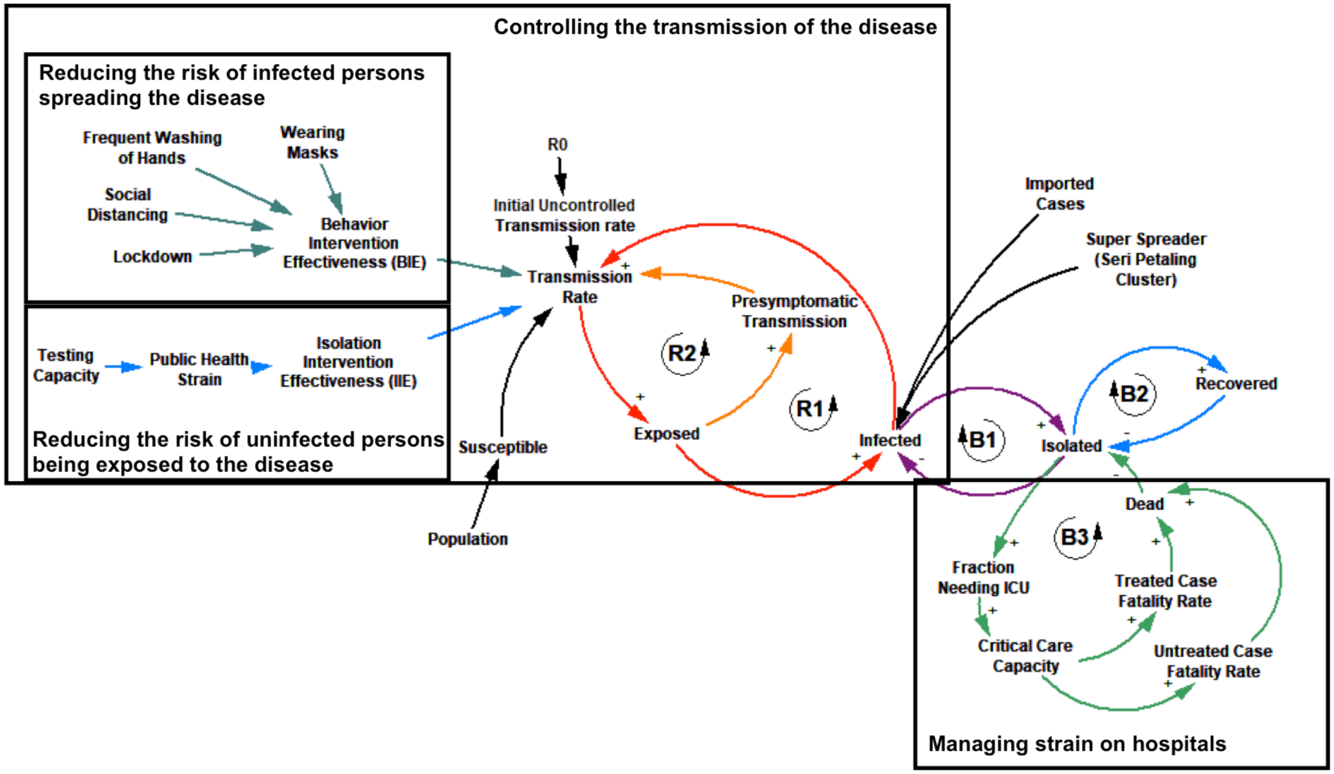


Figure S3. Causal loop diagram of augmented SEIR Model. The causal loop diagram shows the major relationships (links) between the parameters in two sub-systems namely: the transmission rate and the strain on hospitals. The transmission rate is calculated based on parameters that reduce the risk of infected persons spreading the disease and the risk of uninfected persons being exposed to the disease. The strain on hospitals is based on critical care capacity, namely the number of intensive care (ICU) beds.
